# RELATIONSHIP BETWEEN HELMINTHASIS AND GASTRIC CANCER: A SYSTEMATIC REVIEW

**DOI:** 10.1101/2021.08.07.21261231

**Authors:** David Fabian Ramírez Moreno

## Abstract

**Introduction:** helminths are parasitic worms able to produce diverse clinical manifestations in humans, mainly in the gut. Gastric cancer it’s a high incidence entity in Colombia, being the highland regions where it’s incidence is the highest in comparison with the lower incidence coastal region. From the above it is intended to determine the relationship between helminthiasis and the development of gastric cancer.

**Methodology:** A systematic review was performed in four databases for studies evaluating the relationship between helminthiasis and the development of gastric cancer.

**Results:** We included 16 articles from 929 records, with 11 articles reporting a positive relationship and 5 articles with negative relationship.

**Conclusions:** Parasitic infections of the gastrointestinal tract by helminths promote TH-2 type immune responses and decrease TH-1 type that are involved in the progression of precancerous lesions associated with *Helicobacter pylori* infection.

## INTRODUCTION

Gastric cancer is known to be a pathology of high morbidity and mortality worldwide, ranking as the second type of cancer that causes the most deaths in the world (1), and the fourth cause of new cancer cases per year. According to reports from the year 2000 and 2013, 945,000 (2) and 984,000 new cases respectively are reported worldwide, with 841,000 deaths (3). Japan, China, Eastern European countries and tropical South American countries are some of the places with low rates of this, while the United States, Australia and Africa are some of the places with low prevalence and incidence of gastric cancer, having an adjusted world rate of 15.62 x 100,000 inhabitants (4). The variations with respect to sex are not significant, describing a proportion twice as frequent in men than in women. The highest incidence by age is between 50 and 70 years, with an average maximum incidence around 60 years, being infrequent before 30 years (5). Most of the time, carcinoma of the stomach is in an advanced progressive phase at the time of diagnosis, with infiltration beyond the submucosa and significant invasion of the gastric wall (6).

In Colombia, gastric cancer has been a disease of high prevalence and high morbidity and mortality, being the main cause of death from cancer in both sexes, mainly represented by intestinal-type aden ocarcinoma (7). An estimated incidence of gastric cancer of 20.7 x 100,000 inhabitants was reported for women and men during 1995 and 1999 (2). The incidence for the year 2006 was estimated between 35.62 and 36.78 per 100 000 inhabitants in men, occupying the second place in frequency after prostate cancer. In women, the incidence was found between 22.62 and 24.31 per 100 000 inhabitants, occupying the fourth place in frequency after breast and cervical cancer (8). Between the period 2000-2006 showed about 70 887 annual cases of cancer gastric where the departments of Nariño, Boyacá, Cundinamarca, Tolima, Bogota, Santander, Antioquia, Valle and Norte de Santander reported ro n greater number of cases. (9) Gastric cancer in young patients has a slightly higher prevalence in the country than that reported worldwide, these patients present with a more advanced stage than older patients, presenting mostly diffuse cancer (90 %) which has generated high mortality figures (10).

There are 3 factors that have been studied and mainly associated with the development of gastric cancer: the external environment, predisposition of each individual and the infectious agent *H. pylori*, which together with the presence of risk factors (excessive consumption of salt, overcrowding, consumption of non-potable water and an unbalanced diet) determine the evolution of a normal gastric mucosa until its progressive damage or injury with the subsequent development of atrophic gastritis, metaplasia, dysplasia and gastric adenocarcinoma (1). However, there are few studies that have been done to identify new relationships between the development of adenocarcinoma and external agents, such as that given, for example, with other types of infectious agents.

In 1992, Ho lcombe (11) spoke about the “African Enigma”, in which he described a particularly low incidence rate of gastric cancer in the general African population, coming to relate it to the low prevalence of pre-existing lesions in these individuals, mainly metaplasia, a factor that, according to the author, could constitute a significant risk in patients with *H. pylori infection*. Whary et al (12) and Correa et al (1) reported that *H. pylori infection* and the development of gastric cancer are very common in Colombia, however, in those areas where there was a high prevalence of helminthiasis, the development of of cancer was present to a lesser extent, while Yang et al also speak of possible differences in the genotypic characteristics of the different strains of *H. pylori* present in Colombian populations, which can greatly influence this variation of prevalence (13). Gonzales et al described that *H. pylori* coinfection with certain helminths or other types of parasites can balance this response towards the humoral type (Th2), an effect that in turn reduces the risk of severe and progressive diseases (14) ; On the other hand, studies such as the one carried out by Najjari et al (15) show that there are helminths that can aggravate the inflammatory response, a factor that determines the evolution of gastric cancer.

Helminths are parasitic worms that can live in their host either as larvae or as adults, capable of causing various clinical conditions in humans, mainly of the intestinal type and which in turn are subdivided into two large groups: flatworms (flatworms) and roundworms (roundworms), which differ in different aspects such as their evolution and development, being the most evolved roundworms, having a more complex biological and physiological conformation, in addition to their form and type of reproduction (16). The nematodes are worms elongated cylindrically shaped bilaterally symmetrical with the smaller diameter ends having a separate full, reproductive system developed digestive system and sexes, whose internal organs are contained within a pseudocoel limited externally by a wall formed cuticle, h i podermis and muscular layer (17). Among the most important we have *Ascaris lumbricoides, Trichuris trichiura, Ancylostoma duodenale* and *Necator americanus*, which make up the Uncinarias, *Strongyloides stercoralis*, and *Enterobius vermicularis* (17). As for the flatworms, these in turn are divided into tapeworms and trematodes ; respect to cestodes as important pathogens have to *Taenia solium* and *S aginata, Hymenolepis nana*, among the trematodes to *Fasciolopsis buski* (17).

Helminths are distributed throughout the world and can bring complications from mild to life-threatening. Thus, although many of the diseases they cause will respond to drug treatments, there is the possibility of producing mechanical intestinal obstruction, gastrointestinal bleeding, pancreatitis, and appendicitis, so necessitating the use of techniques surgery for removal of the digestive tract (18), there are even cases in which they are associated with the development of tumors (19).

The main mechanism of transmission of these parasites is through an anus-hand-mouth cycle, which means that their transmission can occur through the consumption of contaminated food or water, an important mechanism in children. Thus, helminthiases are associated with socio-economic and environmental conditions, and can be defined as an indicator of the quality of life and hygiene to which a certain population may be subjected (20). by identifying the various forms of these in fecal or organic samples (21).

Taking into account the above, for the present study, a systematic review was carried out in different databases, with the aim of knowing what type of relationship is between gastric cancer and helminth infections, as well as whether there are differences between the outcome in which a certain type of helminth influences and which of them are more prevalently related.

## METHODOLOGY

A systematic review was carried out with the aim of evaluating the relationship of helminthiasis in the development of gastric cancer, following the recommendations of the *Preferred Reporting Items for Systematic Reviews and Metaanalyses* (PRISMA) *guide*.

### Identification of studies and search strategies

s and sought gastric cancer studies or quad ros predisposing clinical program for you, in patients diagnosed with helminths. A search was made in 4 multidisciplinary databases: Pub-Med, Embase, EBSCO and LILACS, from the year 2000 to 2017, using the following search scheme: [stomach neoplasm OR gastric Cancer OR gastric neoplasm] AND [Helminthiasis]. Additional filters were not taken into account, except for the language limitation to Spanish and English.

### Screening or application of inclusion criteria

1) Investigations whose subjects were infected by a helminth type parasite and the diagnosis was made by means of a laboratory test ; 2) Research where a relationship between helminthiasis and the development of gastric cancer was described or demonstrated; 3) Original experimental articles type clinical trial, cases and controls and cohort studies.

### Choice or application of exclusion criteria

1) Articles published before the year 2000; 2) Articles with a different theme from that proposed in the study, review articles, meta-analysis, letters to the editor, case reports and opinion reports; 3) Investigations in which there is any factor that could alter or mask the direct action of the helminth against the development of gastric cancer.

### Collection of information

P ara search, was assigned to each researcher, randomly, a database for an independent evaluation. All the records resulting from the search were evaluated, titles and abstracts were recorded, evaluating the inclusion and exclusion criteria. The full text of the selected studies was obtained and another reviewer re-verified the fulfillment of these criteria.

The main variables of the study were the relationship between helminthiasis and the development of gastric cancer, defined as a risk factor, a protective factor or a factor with little significant relationship. The secondary variables were: year of publication, type of study subject, parasite involved, interrelation with the development of gastric cancer, and type of study design.

## RESULTS

The relationship between helminthiasis and the development of gastric cancer was evaluated in 16 studies (Graph 1), within which the appearance of different pathological changes in the gastrointestinal epithelium in relation to the presence of some helminth was discussed, both experimentally, as retrospective and observational.

**Figure 1.**
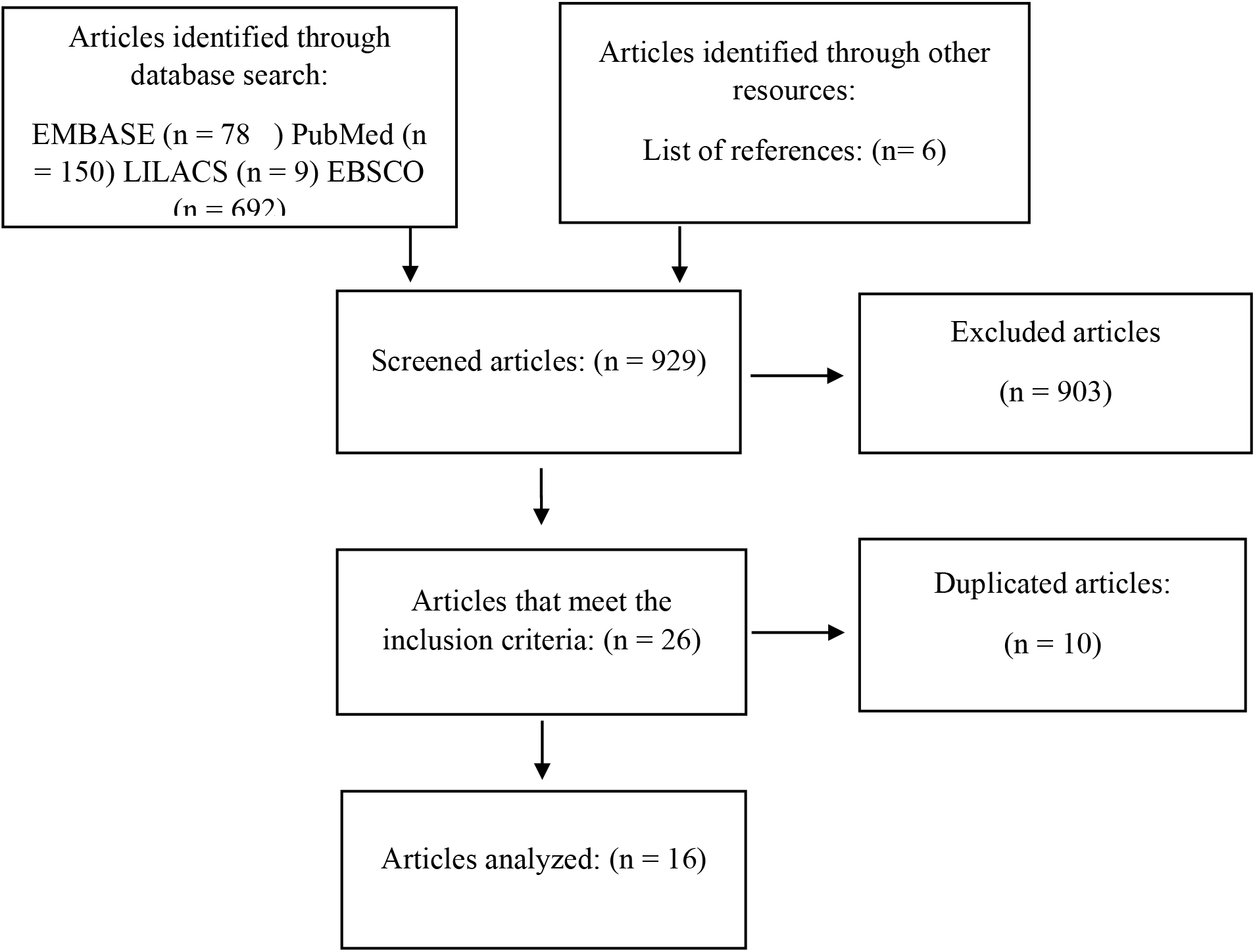
Search algorithm.

The established relationship could be classified into a positive interaction (helminthiasis as a protective factor against gastric cancer), negative (helminthiasis as a risk factor against gastric cancer) or null (there is no significant relationship between helminthiasis and the development of gastric cancer). Most of the studies were evaluated in the human population, except for Martin et al. (22) and Sargar et al. (23), who performed it on gerbils; and Najjar et al. (15), Fox et al., Whary et al. (12) And Kara et al. (24), who performed them in mice with different characteristics.

Studies were assessed according to Cochrane guidelines. The studies do not specify the existence or method of randomization for evaluated groups and control groups. The available abstracts of each of the studies were included for selection for the establishment of their inclusion or exclusion, in which only the information on the main outcome was presented. Therefore, the studies may present bias in the selection and information.

### Positive relationship

In total, 11 articles elucidated a positive relationship, 6 in humans and 5 in animals, defining it as the establishment of helminthiasis as a protective factor in the evolution of the disease, macroscopic and microscopic pathological characteristics and outcome in a certain time. on the development of l gastric cancer or estadi or history of the same (gastri tis, atrophic gastritis, metapla sia, ulcers appearance, nodules and lesions in both the epithelium and in gastric mucosa). The immune response may play a fundamental role in the regulation of pathological evolution in subjects infected with *H*. *pylori* through modulation of the TH2 response, mainly in coinfections with filariae (22) and *Trichinella spiralis* (23), which leads to an increase in the production of factors such as IL-10 and interferon gamma, protecting the epithelium and the gastrointestinal mucosa from the harmful effects of the inflammatory response generated by TH1 (25). In studies with *Heligmosomoides polygyrus*, it was shown that, although mice infected by the parasite develop chronic inflammation with the appearance of foveolar hyperplasia and early dysplasia and mice infected with *H. pylori* non-invasive neoplasms and intramucosal carcinomas, in coinfection, a significant reduction was evidenced gastric atrophy, dysplasia and none developed neoplasms (12,26,27), verifying that in coinfected rodents, there is a significant TH2 response with subsequent release of high levels of IL-10, IL-4, interferon gamma and activation of lymphocytes both CD4 + and CD48 +, which in turn increase IgG production by inhibiting IgE (24).

In studies in human populations, it has been shown that the presence of *H. pylori* is related to a significant increase in lipid peroxidation and nitric oxide, as well as a reduction in anti-oxidant agents (SOD, CAT, GSH), the opposite being the case in the presence of a coinfection with *Schistosoma mansoni* (28) that would seem to modulate the inflammatory response against infection by the bacteria, reducing DNA damage, apoptosis, cell proliferation and the increase of anti oxidants, inferring a complex interaction between the responses immunological TH1, TH2 and Treg (29). In infections caused by *Ascaris lumbricoides* and *Trichuris trichiura*, a strong Th2-type immune response was also evidenced, which in turn leads to the high production of IL-10 and growth factor B, controlling inflammatory reactions by suppressing the action of other cytokines (30). Other studies of coinfections with *Schitosoma japonicum* and *H. pylori*, in feces show that the concurrent infection of *S. japonicum* modifies the serological responses of IgG to *H. pylori*, reducing the probability of developing gastric atrophy (31). The impact of infection with the intestinal helminths *A*. *lumbricoides* and *T*. *trichiura* was investigated by Geiger et al. (32) in young adults, where the specific cellular response capacity was greater in parasite-free endemic controls than in patients infected with *T. trichiura, A. lumbricoides*, or patients coinfected with both parasites. In Colombian studies, when comparing different geographic populations, Whary et al. (12), inferred that parasitic infections in children could polarize the TH1 response to TH2 and therefore alter the immune reaction in *H. pylori infection* ; The difference in the development of gastric cancer in different regions of Colombia compared to the presence of intestinal helminth infections was also demonstrated, detecting a significant association between the seropositivity of *A. lumbricoides* and a greater response of TH2 / IgG1 to *H. pylori*, thus presenting helminthiasis as a protective factor against the development of this cancer (33).

### Negative relationship

E n total, 5 items concluded a negative relationship, four human and one animal, defined the same as the establishment of helminthiasis as a risk factor in development of the disease, pathological features macroscopic and microscopic and outcome in certain time on the development of gastric cancer or some determining stage of it. Studies were performed with mice infected with *Hysterthylacium amoyense*, Find ndose in the group evaluated that 3 of 5 mice infected with the parasite developed tumor nodules which would be diagnosed with cancer, rel to probably amateurs antigen excretory-secretory (ES), active in the recruitment of eosinophils, which would worsen the inflammatory response (15). It is suggested that the chronic inflammation caused by the parasite, and a possible tumorigenic effect of certain Anisakis parasitic secretions can produce local persistent inflammation and granulomas, in addition larvae have been found in gastrointestinal tumors, having evidence of a significant negative relationship (34). In studies on infection by *Strongyloides stercolaris*, a notable inflammatory infiltrate was found in all, development of glandular atrophy with type 2 intestinal metaplasia and gastric adenocarcinomas (35). Waku et to the evaluated 1,199 people who had schistosomiasis, amebiasis, tuberculosis and candidiasis, and concluded that there is a relationship between the generated infections by these pathogens and development of neoplasms of the gastrointestinal tract.

**Table 1.** Characteristics of the articles included.

| Author | Year | Country | intervention (helminthiasis ) | Relations hip Type | Study populatio n | Amount studied | Results |
| --- | --- | --- | --- | --- | --- | --- | --- |
| Whary | 2014 | USES | Yes, *Heligmos omoides polygyrus* | Positive | Mice | - | In mice with coinfection by *Heligmosomoides polygyrus* a nd *Helicobacter pylori* , gas atrophy , dysplasia and development of neoplasms caused by immune processes against these pathogens was lower compared to monoinfected mice . Not only inflammation influences carcinogenesis by *H.pylori* , bacterial flora may be involved in generating changes in pH. |
| Whary | 2005 | Colombia | Yes | Positive | Humans | 454 | Infections helmintiá SICAS in children promotes Th2 immune response type with increased levels of IgE and IgG1 that may decrease the risk of gastric cancer in these individuals later in life. |
| Fox | 2000 | USES | Yes *Heligmosomoides polygyrus* | Positive | Mice | - | In coinfection *Helicobacter felis* and *Heligmosomoides polygyrus* (compared to mice monoinfection by *H.pylori* ), gastric atrophy associated with *H.pylori* was considerably reduced despite the high and chronic inflammation colonization by the bacteria, that associated with decreased in mRNA for cytokines and chemokines responsible for the Th1 immune response. |
| Waku | 2005 | Italy | Yes | Negative | Humans | 1199 | Helminth infection produces an inflammatory response leading to the development of gastro i tis atrophic. |
| Elshal | 2004 | Egypt | If<br>*Schistosoma mansoni* | Positive | Humans | 73 | The coinfection by *Schistosoma mansoni* and *H. pylori* is associated with gastritis reduced incidence of 1 etaplas to the modular *S. mansoni* the inflammatory response, by a Th2 - type response. |
| Figuereido | 2009 | Brazil | If<br>*Trichuris trichura* and *Ascaris lumbricoides* | Positive | Humans | 1445 | The immune response generated helminthiasis TH2 - type leading to a high production of IL10 and factor I growth B, controlling re to inflammatory ctions to suppress the action of the other citocinas. |
| Yiqi DU | 2006 | China | If<br>*Schistosoma japonicum* | Positive | Humans | 150 | The coinfection by *Schistosoma japonicum* modulates TH1 response *H. pylori* increasing the TH2 response with increased levels of IL-10, IgG1 and IgE . |
| Filbey | 2014 | Amsterdam | If<br>*Heligmosomoides polygyrus* | Positive | Mice | - | The helminth-mediated TH2 response increases the production of IL 10, IgG and decreases the production of IgE . |
| Courtney | 2012 | USES | Yes<br>*Ascaris lumbricoides* | Positive | Humans | 240 | The prevalence of gastric cancer in Tumaco is lower compared to areas of high altitudes such as Pasto / Tuquerres , while the prevalence of 1 stage 1 s is higher in Tumaco. *Ascaris lumbricoides* infection generates a great Th2 / IgG1 response against *H. pylori* infection , creating protection for cancer caused by this bacterium. |
| Cherian | 2010 | Australia | Yes | Positive | Humans |  | In *H. pylori* and helminth infection , there is an elevation in the production of IL-10 and IL-12, as well as a complex interaction between Th1, Th2 and Treg immune responses . |
| Sagar | 2004 | Canada | Yes<br>*Trichinella spiralis* | Positive | Gerbils | - | *Trichinella spiralis* produces a modulation of the TH2 response , with an increase in the levels of IL 10 and IgG1 |
| Najjari | 2016 | Iran | Yes<br>*Hysterothylacium amoyense* | Negative | Mice | 10 | *Hysterothylacium amoyense* associated with the appearance of tumor nodules in infected mice. |
| Geiger | 2002 | Germany | Yes<br>*Ascaris lumbricoides* | Positive | Humans | 579 | *Ascaris lumbricoides* infection modulates the Th1-type immune response, reducing it and generating a mainly Th2 response that at the same time acts as a protective mechanism against an autoimmune reaction that generates pathological damage. |
| Garcia | 2015. | Spain | Yes<br>*Anisakis spp.* | Negative | Humans | 94 | Infections with *Anisakis spp.*. They can produce persistent inflammation and gastrointestinal tumors. |
| Rivasi | 2006 | Italy | Yes<br>*Strongyloides stercoralis* | Negative | Humans | fifteen | Subjects who showed symptoms of peptic -péptica and infected by *Strongyloides stercoralis* had infiltrated inflammation, 1 etaplasia intestinal and gastric adenocarcinoma in some cases, being elevated Th2 type immune, causing pathological changes response. |
| Martin | 2010 | USES | Yes<br>*Brugia pahangi* | Positive | Gerbils | 85 | After 21 weeks, a significant change was evidenced in terms of reducing pathological changes in mice coinfectad with *Brugia pahangi* and *H. pylori*. |

## DISCUSSION

The objective of this study was to investigate the effect on the immune response in subjects infected with helminths, its possible relationship with gastric cancer or its progression from predecessor stages of the disease. It has been described that when infected with *H. pylori* bacterial antigens such as the B subunit of urease (36) are recognized by pattern recognition receptors that will initiate the immune response (37) this response is characterized by a process called immunological polarization (38), which consists of processes that are performed for one of the two predominant types of lymphocytes helper 1 and 2 (Th1 and Th2) that come from the same precursor, the T lymphocyte naïve (Th0) is Diff and to one of the two lines predominantly, polarization occurs to the profile of cyto c inas to the display above, the most important are interleukin 12 (IL-12) and interleukin 10 (IL-10), if the cells Th0 are exposed s to high levels of IL-12 and Th1 differentiate directly inhib would production and development of Th2 cells, by secreting molecules as TIM-3 (37,39,40) with a response predominantly of T helper cells 1 (Th1), which mediate the cellular immune response, favor tissue damage and inflammation through different mechanisms that include the production of molecules such as interferon gamma (INF-γ) and tumor necrosis factor alpha (TNF-α) (41), these molecules cause the release of IL-8 in the epithelial cells of the gastric mucosa, which is a potent neutrophil chemotactic factor (41), promote tissue damage and cause an increase in the expression of proteins of the major complex of histocompatibility (HCM) that could favor *H*.*pylori* infection (42). This phenomenon also occurs when an individual is infected with intestinal helminths (43), the intestinal epithelium cytosines released as the s interleukin s 10, 25 and 32 having function alarminas quickly initiate polarization towards Th2 and molecules as lymphopoietin stromal thymic which has a function similar to TIM-3 but suppressing the TH1 response (1.44). D and so hypothetically a polarization towards Th2 decrease adverse events of cellular immunity mediated by Th1 cells towards the gastric mucosa (45). This hypothesis has been evidenced in studies in the murine model where the expression of TIM-3 is associated with a worse prognosis in gastric cancer (46) and experimental studies in which molecules that block this factor are used to prevent its progression (46).

In the review studies which found in individuals with predisposing factors for gastric cancer, usually acute or chronic infection with *H. pylori* in co -infections with helminths such as heartworms, *Trichinella spiralis, Heligmosomoides polygyrus, Schistosoma mansoni, Asc aris lumbricoides, Trichuris trichiura, S. japonicum*, presented elevated levels of IL-10, IL-4 and immunoglobulin G subtype 1 (IgG1), factors that would polarize the immune response towards Th2, this together with lower levels of gastric cancer development than their non-parasitized counterparts, which which contributes to provide evidence that would support the theory described above. As a counterpart, descriptions of helminths that could be involved in the development and severity of gastric cancer were found, such as: with *Hysterthylacium amoyense, Anisakis spp, Strongyloides stercolaris*, the damage produced by these helminths is apparently by products secreted by them. *Strongyloides stercolaris* also seems to be related to lymphomas and a case has been reported in which it is associated with gastric adenocarcinoma (20, 21).

In this review, points have also been found in which contradiction is evident. Figuereido et al. (30) report that the transforming growth factor beta TGF-β would be one of the factors that modulates the inflammatory response, decreasing the damage and progression to gastric cancer, in contrast to this later articles that report that TGF-β favors invasion and metastasis cancer cells (49–52). Additionally, in the articles included in the review, schistosomiasis is presented as a positive (28,31) and negative (27) factor for the development of gastric cancer, this parasitosis has also been associated with bladder cancer (53,54), therefore that more research is needed on how this helminth might or might not cause cancer.

Other publications have reported results that contradict what is said in this study demonstrate protection against *H. pylori* at the acti v ation of Th1 (36) but without comparing response to helminth intestinal.

In conclusion, some helminths producing parasitism in humans can generate stimuli that polarizes n response immune cell, proinflammatory Th1 humoral Th2 by the secretion of a specific profile of cytokines may thus have a positive impact on the gastric lesion and progression gastric cancer; without however, other helminths may in turn produce a chronic inflammatory response participating as an important factor in the pathophysiology neoplastic

## Data Availability

All of the data supporting the results of this paper is reported on the paper itself.

